# Sleep in Frontline Healthcare Workers on Social Media During the COVID-19 Pandemic

**DOI:** 10.1101/2021.01.19.21250128

**Authors:** Nancy H Stewart, Anya L Koza, Serena Dhaon, Christiana Shoushtari, Maylyn Martinez, Vineet M Arora

## Abstract

**Importance:** During the pandemic, healthcare workers on social media are sharing their challenges, including sleep disturbances.

**Objective:** To assess sleep using validated measures among frontline healthcare workers on social media

**Design:** A self-selection survey was distributed on Facebook, Twitter, and Instagram for 16 days (August 31-September 15, 2020) targeting healthcare workers (HCW) who were clinically active during the pandemic. Study participants completed the Pittsburgh Sleep Quality Index (PSQI), Insomnia Severity Index (ISI), and reported demographic/career information. Poor sleep quality was defined as PSQI>5. Moderate-to-severe insomnia was defined as an ISI>14. The mini-Z was used to measure burnout. Multivariate logistic regression tested associations between demographics, career characteristics, and sleep outcomes.

**Setting:** Online self-selection survey on social media

**Participants:** 963 surveys were completed. Participants were predominantly White (92.8%), female (73.4%), aged 30-49 (71.9%), and physicians (64.4%). Mean sleep duration was 6.1 (SD 1.2) hours. Nearly 90% reported poor sleep (PSQI). One third (33.0%) reported moderate or severe insomnia. Many (60%) experienced sleep disruptions due to device usage or had bad dreams at least once per week (45%). Over 50% reported burnout. In multivariable logistic regressions, non-physician (OR 2.4; CI: 1.7, 3.4), caring for COVID-19 patients (OR 1.8; CI 1.2, 2.8), Hispanic ethnicity (OR 2.2; CI: 1.4, 3.5), being female (OR 1.6; CI 1.1, 2.4), and having a sleep disorder (OR 4.3; CI 2.7,6.9) were associated with increased odds of insomnia. In open-ended comments (n=310), poor sleep mapped to four categories: children and family, work demands, personal health, and pandemic-related sleep disturbances.

**Conclusion:** During the COVID-19 pandemic, 90% of frontline healthcare workers surveyed on social media reported poor sleep, over one-third reported insomnia, and over half reported burnout. Many also reported sleep disruptions due to device usage and nightmares. Sleep interventions for frontline healthcare workers are urgently needed.

**Key points:** *Question:* How are frontline healthcare workers on social media sleeping during the pandemic?

*Findings:* During the COVID-19 pandemic, 90% of frontline healthcare workers on social media are reporting poor sleep, and one third are reporting insomnia. Those who report sleep disturbances were more likely to report burnout.

*Meaning:* Interventions aimed at improving the sleep of frontline healthcare workers are warranted.

## Background

Since March 2020, the severe acute respiratory syndrome coronavirus 2, SARS-CoV2 (COVID-19) pandemic has impacted all of our lives on a global scale, especially healthcare workers on the frontlines. During the pandemic, healthcare workers (HCW) have used social to share a variety of concerns relating to their experiences on the frontlines, such as lack of personal protective equipment (PPE), mental anguish, burnout,^1^ and toll caring for such patients. One such concern also includes sleep loss and other sleep disturbances.^2^ While these sleep issues certainly can be attributed to long hours, they may also be due to other reasons. Social media posts suggest that healthcare workers are suffering from bad dreams, insomnia, and also “doomscrolling” - or using mobile devices at night to review social media.^3,4^ Given the increased professional and personal responsibilities for healthcare workers during the pandemic, it’s possible that sleep loss or disturbances could also be compounded by additional worries during the pandemic. Moreover, there is mounting evidence demonstrating inadequate physician sleep is associated with burnout and medical errors.^5,6^

Several studies in other countries describe the sleep of HCW during the COVID-19 pandemic. Most notable are two meta-analyses on sleep in HCW during the initial months of the COVID-19 pandemic, with studies mostly focusing on the experience of HCW in China. Pappa et al. ^7^ reviewed 5 cross-sectional studies conducted among Chinese HCW prior to April 17, 2020 and reported a prevalence of insomnia (38.9%) among them. Salari et al.^8^ reviewed 7 cross-sectional studies from Asia and the Middle East conducted among nurses and physicians prior to June 24, 2020 and reported a prevalence of sleep disturbances of 34.8% and 41.6% in nurses and physicians, respectively. Additionally, several other studies reported on sleep loss in HCW from countries such as Oman,^9^ Bahrain,^10^ and Spain.^11^

To date, there have been few studies of the sleep experience among frontline HCW in the United States (U.S.). These studies did not utilize validated measures of sleep and were limited to a single institution.^3,12^ In the U.S., to our knowledge, no study has yet to evaluate sleep using standardized measures in clinically active frontline HCW during the COVID-19 pandemic. Despite anecdotal observations of sleep loss described via social media platforms, no study yet has explicitly explored the sleep of frontline HCW in the COVID-19 pandemic utilizing social media, an increasingly important and influential voice during the pandemic. This study aims to describe the sleep disturbances of U.S. frontline HCW on social media during the COVID-19 pandemic using validated instruments. The study also aims to examine the association between various demographic and career characteristics of HCW with reported sleep disturbances and the association between sleep disturbances and burnout.

## Methods

From August 31 to September 15, 2020, we recruited frontline HCW from across the world to complete a web-based R.E.S.T. (Recommending Essential Sleep Time) Survey. The restricted, self-selection survey was distributed through Twitter, Facebook, and Instagram social media accounts made specifically for the study. A river sampling method was used to recruit subjects anonymously.^13^ This online sampling technique redirected interested subjects to the survey link via digital promotion (e.g., banners, advertisements, flyers, offers).^13^ A unique “bit.ly” link was created for the social media accounts to enable tracking of views and clicks. Participation in the study was voluntary, and participants could opt-out at any time while responding to survey questions. Subjects included physicians, physician assistants, nurse practitioners, nurses, pharmacists, physical/occupational/respiratory therapists, healthcare trainees, and allied HCW (e.g., medical technologist, therapist, certified nursing assistant). The University of Chicago Institutional Review Board reviewed and approved the protocol.

### Survey Design

Study data were collected and managed using REDCap (Research Electronic Data Capture) hosted at the University of Chicago. REDCap is a secure, web-based software platform that supports data capture for research studies.^14,15^ Qualifying participants were at least 18 years of age and frontline HCW clinically active during COVID-19. The survey automatically terminated for those that did not meet these inclusion criteria. We asked subjects to frame all their responses for a month in the pandemic where they experienced the greatest clinical intensity and risk of COVID-19 transmission.

The survey contained the following categories: sleep, well-being, career, and baseline demographics. The *Sleep* section included the Pittsburgh Sleep Quality Index (PSQI)^16^ and the Insomnia Severity Index (ISI).^17^ The PSQI is a 19-item validated questionnaire that measures sleep quality over one month, generating one global score.^16^ The ISI is a 7-item validated questionnaire that assesses the nature, severity, and impact of insomnia in adults.^17^ The *Sleep* section also asked about sleep disturbances due to device usage. The *Well-being* section included the Mini-Z Burnout Survey, an 8-item validated questionnaire that investigates healthcare burnout and job satisfaction.^18^ The Mini-Z also includes the 1-item burnout measure that is validated against the longer Maslach Burnout Inventory.^19^ Other questions in this section related to pandemic-related worries, changing responsibilities, living situation, and health status. The *Career* section collected data on the subject’s job role, level of training, specialty, and career setting prior to the pandemic and whether they cared for COVID-19 patients directly. Baseline demographics were collected on marital status, age, gender, race, ethnicity, type of practice, location of practice, and the type of community served (e.g., urban, suburban, rural, etc.). Two open-ended questions were also included in the survey: “What was the cause of your sleep disruption(s) during your reported month?” and “Is there anything else you would like to share?”

Upon completion of the survey, participants were invited to submit their email address for a chance to receive daily raffled $25 Amazon gift cards (14 gift cards in total). Before deployment, the survey was piloted with a few HCW not affiliated with the study. Changes made included deleting duplicate questions and removing one sleep scale to abbreviate the survey to 10 minutes or less.

### Social Media Distribution Strategy

To disseminate the survey link, we created a Facebook page and a Twitter and Instagram account on behalf of the R.E.S.T. study. This allowed the use of engagement tracking built into these social media platforms to calculate the response rate. To market our survey through Instagram, we used paid methods of advertising. On Facebook, we posted on healthcare-specific social media groups (e.g., Women Physicians Wellness, StyleMD, and the Physician Collective). On Twitter, we tagged colleagues and organizations and employed healthcare-specific hashtags (e.g., #MedTwitter, #NurseTwitter). While the authors re-shared the R.E.S.T. Survey’s posts on their personal Twitter accounts to raise visibility, the link remained connected to the R.E.S.T. accounts’ original posts to preserve the ability to track engagement. To promote the survey, an infographic was posted each day on the study’s social media accounts. Survey links were also placed in each social media accounts’ biography. To draw attention to the survey post and the account, we linked each post to standard information from national organizations (e.g., American Academy of Sleep Medicine and the Sleep Research Society) and to articles about sleep and the pandemic through the study’s social media accounts.

One of our study authors (SRD) used a customized *bit*.*ly* link (bit.ly/RESTsurvey) to track user engagement daily on Facebook, Twitter, Instagram, and email. The metrics analytics from the social media platforms enabled us to view specific metrics on reach (the total number of individuals who view the content) and link clicks (the number of times individuals choose to open the survey). At midnight of each day, we calculated the study’s reach and link clicks on each social media website. This allowed us to compare the performance of each platform and track our progress. We also calculated the number of survey responses per day while the survey was active.

### Statistical Analysis

Descriptive statistics were used to quantify demographics and profession. Primary outcomes were poor sleep quality and moderate-to-severe insomnia. Poor sleep quality was defined as a PSQI score > 5.^16^ Moderate-to-severe insomnia was defined as an ISI score of >14.^17^ Mini-Z was used to measure burnout.^18^ Multivariate logistic regression was used to test for independent associations between age, gender, race (Black vs. non-Black), ethnicity (Hispanic vs. Non-Hispanic), and profession (physician vs. non-physician), with odds of each of the outcomes, after controlling for those with a preexisting sleep disorder. All statistical analyses were conducted using StataCorp. 2019. Stata Statistical Software: Release 16. College Station TX: StataCorp LLC with P< 0.05 used to indicate statistical significance.

### Coding Open-Ended Comments

There were two open-ended questions in the survey. For those that responded to one or both questions, open coding of comments was utilized.^20^ Sleep disturbance mapped to four main themes: (1) Children and family; (2) Work demands affecting sleep; (3) Personal health conditions; and (4) Pandemic-related sleep disturbances. Sub-themes were created for each.

## Results

Our social media posts were seen by 87,061 unique individuals and resulted in 976 clicks on our survey link. Our final sample had 963 surveys that were submitted. The most common ways people discovered the survey were through Twitter (39.4%), private Facebook group (21.1%), and a colleague (10.9%).

Participants were mostly female (73.4%), White (92.8%), aged 30-49 (71.9%), and physicians (64.4%). (Table 1)

**Table 1.** Sample Respondent Characteristics (SLEEP, using ISI p)

|  | N (%) |
| --- | --- |
| Total | 963 (100) |
| Age Group |  |
| 18-29 | 149 (15.5) |
| 30-49 | 692 (71.9) |
| 50+ | 86 (8.9) |
| Did not answer | 36 (3.7) |
| Gender |  |
| Male | 220 (22.9) |
| Female | 707 (73.4) |
| Did not answer | 36 (3.7) |
| Race |  |
| White | 894 (92.8) |
| Black | 69 (7.2) |
| Ethnicity |  |
| Non-Hispanic | 759 (78.8) |
| Hispanic | 124 (12.9) |
| Did not answer | 80 (8.3) |
| Significant Other |  |
| Yes | 700 (72.7) |
| No | 220 (22.9) |
| Did not answer | 43 (4.5) |
| Career Role |  |
| Physician | 620 (64.4) |
| Non-physician | 343 (35.6) |
| Directly Cared for COVID-19 patients | 744 (80) |
| Country/Continent |  |
| -United States | 882 (91.6) |
| - Canada | 18 (1.9) |
| -Central America | 10 (1.0) |
| - South America | 9 (0.9) |
| -Europe | 7 (0.7) |
| -Asia/Oceania | 4 (0.4) |
| -Australia/New Zealand | 4 (0.4) |
| Africa | 3 (0.3) |
| Did not answer | 26 (2.7) |

Poor sleep quality, defined as a PSQI score > 5, was identified in 88.9% (n = 753) of all healthcare workers (Table 2). Rates of moderate and/or severe insomnia were 28.5% and 4.5%, respectively (Table 2). Average sleep duration was 6.1 +/- 1.2 hours (Table 2). Many (60%) reported sleep disruptions due to device usage or due to bad dreams at least once per week (45%). Those with a sleep disorder were more likely to report burnout, 52.1% vs 63.2% [x2 (4, N= 963) = 116.4, p < 0.001)]. In unadjusted analyses Black race (31.0% vs 59.4%), Hispanic ethnicity (27.9% vs 59.6%), being single (44.6% vs 27.1%), and being a non-physician (50.7% vs. 23.2%) were significantly associated with risk of moderate-to-severe insomnia.

**Table 2.** Prevalence of Poor Sleep and Insomnia in Healthcare Workers Working During the COVID-19 Pandemic

|  | N (%) |
| --- | --- |
| Poor Sleep Quality<sup>a</sup> | 963 (90.1) |
| Insomnia<sup>b</sup> |  |
| Absent | 220 (25.1) |
| Sub-threshold | 367 (41.9) |
| Moderate | 249 (28.5) |
| Severe | 39 (4.5) |
| Average Sleep Duration (hours) | 6.1 +/- 1.2 |

<sup>a</sup> As measured by the Pittsburgh Sleep Quality Index (PSQI); Poor sleep quality was defined as PSQI score >5.<sup>16</sup>
<sup>b</sup>As measured by the Insomnia Severity Index (ISI); Insomnia qualified by ISI scores and interpreted as absent (0-7), subthreshold (8-14), moderate (15-21), or severe (22-28).<sup>17</sup>
Average hours of sleep duration was defined by the PSQI.<sup>16</sup>
<sup>c</sup>Burnout measured by the one item Mini-Z Burnout survey

In multivariable logistic regressions, non-physician (OR 2.4; CI: 1.7, 3.4), caring for COVID-19 patients (OR 1.8; CI 1.2, 2.8), Hispanic ethnicity (OR 2.2; CI: 1.4, 3.5), and being female (OR 1.6; CI 1.1, 2.4) were associated with increased odds of insomnia (Table 3). Having a sleeping disorder also increased odds of insomnia compared to those without one (OR 4.3; CI 2.7,6.9). (Table 3).

**Table 3.** Risk Factors for Insomnia* Among Frontline Healthcare Workers During the COVID-19 Pandemic

| N = 963 | Insomnia<sup>a</sup><br>(OR, CI) | p-value |
| --- | --- | --- |
| Age Group |  |  |
| 18-29 | Ref |  |
| 30-49 | 1.1 (0.7, 1.8) | 0.63 |
| 50+ | 1.3 (0.7, 2.4) | 0.49 |
| Gender |  |  |
| Male | Ref |  |
| Female | 1.6 (1.1, 2.4) | < 0.05 |
| Race |  |  |
| Black | 1.5 (0.8, 2.7) | 0.24 |
| Ethnicity |  |  |
| Hispanic | 2.2 (1.4, 3.5) | < 0.001 |
| Significant Other |  |  |
| No | 1.3 (0.9, 1.9) | 0.18 |
| Career Role |  |  |
| Physician | Ref |  |
| Non-physician | 2.4 (1.7, 3.4) | < 0.001 |
| Prior Sleep Disorder |  |  |
| Yes | 4.3 (2.7, 6.9) | < 0.001 |
| Cared for COVID-19 Patients |  |  |
| Yes | 1.8 (1.2, 2.8) | < 0.01 |

<sup>a</sup> As measured by the Insomnia Sleep Index (ISI); Insomnia defined by scores qualifying as moderate-to-severe insomnia (ISI score >14).<sup>17</sup>
OR = Odds Ratio
CI = 95% Confidence Interval

Three hundred and ten open-ended comments regarding factors affecting sleep were categorized into the following four themes: (1) Children and family; (2) Work demands affecting sleep; (3) Personal health conditions; (4) Pandemic-related sleep disturbances. The most frequently reported non-pandemic-related theme was children and family (n = 59), (“COVID plus home stress plus stress over my kids, my job, my marriage.”) (Table 4). Other non-pandemic related sleep disturbances included work demands affecting sleep (n = 48) (“The volume of calls and messages from my patient and caregiver population is through the roof and I’m sleeping 4-5 hours per night.”) and personal health (n = 41), (“Insomnia predating COVID, but worsened with COVID.”) (Table 4). Pandemic-related sleep disturbances (n = 48), (“I never had sleep issues prior to the COVID-19 pandemic; suddenly I had issues with sleep initiation.”) were also noted (Table 5).

**Table 4.** Categories from Open Ended Comments: Non-Pandemic Related Sleep Disruptions

| Theme | Sub Theme | Example |
| --- | --- | --- |
| Children and Family (n=59) | Childcare (1-18yr) (n=27, 45.8%) | 6 year old who wakes 2-3 times per night (some secondary to COVID related anxiety for him), 2.5 year old who wakes about 2-3 nights per week. |
|  | Infant care (n=18, 30.5%) | Both COVID and my usual sleep disturbances plus my infant daughter. |
|  | Concerns about family affecting sleep (n=8, 13.6%) | COVID plus home stress plus stress over my kids, my job, my marriage. |
|  | Pregnancy affecting sleep (n=6, 10.2%) | I was pregnant with twins and carried to term. COVID increased my anxiety and work-related stress. I had to lay off employees and reduce my salary, right before maternity leave. |
| Work Demands Affecting Sleep (n=48) | Shift work (n=18, 37.5%) | Mostly schedule fluctuations each week - late evening shifts followed by early morning shifts. |
|  | Work Life Interference (n=18, 37.5%) | Extra work at home [charting] due to increased patient volumes. |
|  | Residency Training (n=6, 12.5%) | Switching schedules with residency (days/nights and 28-hour calls), overall sleep debt, and fatigue. |
|  | Volume of calls/paging (n=6, 12.5%) | The volume of calls and messages from my patient and caregiver population is through the roof...sleeping 4-5 hours [per night]. |
| Personal Health Conditions (n=41) | Poor sleep prior to the pandemic (n=18, 43.9%) | I have never been a great sleeper. |
|  | Chronic medical issue (n=12, 29.3%) | Possibly COVID-19 related.... I had an onset of Obsessive-Compulsive Disorder this summer and have been adjusting to antipsychotic medication, intervention, and prescription sleep aids. |
|  | Formal sleep disorder<br>(n=8, 19.5%) | Insomnia pre-dating COVID, but [it] worsened with COVID. |
|  | Female hormones<br>(n=3, 7.3%) | In vitro fertilization side effects. |

**Table 5.** Categories from Open Ended Comments: Sleep Disruptions Due to Pandemic

| Theme | Sub Theme | Example |
| --- | --- | --- |
| Pandemic-Related Sleep Disturbances (n=48) | Pandemic affecting sleep (n=17, 35.4%) | I never had sleep issues prior to the COVID-19 pandemic. Suddenly I had issues with sleep initiation. |
|  | Nightmares about pandemic (n=8, 16.7%) | The only thing that has changed with my sleep is an increase in nightmares and night terrors primarily regarding work situations. |
|  | Pandemic-related exhaustion (n=8, 16.7%) | In the last 1-2 months, I am EXHAUSTED and fall asleep easily, often on the couch right after dinner. |
|  | Medication/Substance use to cope with pandemic (n=7, 14.6%) | Alcohol plays a role in my sleep deprivation and "coping" during COVID. |
|  | Worry about pandemic response (n=4, 8.3%) | I worry about how COVID is being managed by the President of the United States and I don't believe he is doing a good job; I worry things will get even worse if he is elected to a second term. This does keep me awake at night. |
|  | Using electronics to read about pandemic (n=4, 8.3%) | Late night screen time reading about current events. |

## Discussion

To our knowledge, this is the first study to evaluate sleep disturbances among U.S. frontline HCW on social media during the COVID-19 pandemic using self-reported validated measures. Our study demonstrates nearly 90% of surveyed HCW report poor sleep, over 30% report moderate-severe insomnia, and over 50% report burnout. A majority also reported sleep disruptions due to device usage or bad dreams at least once per week. Additionally, survey participants reported a mean sleep duration of 6.1 hours, less than the recommended 7 hours of sleep for U.S. adults.^21^ Controlling for those who had a known sleep disorder, those at highest risk of insomnia included those that were non-physicians, non-male, Hispanic, and those HCW that directly cared for COVID19. We also demonstrate that sleep disturbances in HCW during the pandemic not only relate to work demands and personal health, but also are related to children and family, and to the pandemic itself, including worry about the response to the COVID19 pandemic. Given the strong association between sleep and burnout among health professionals, it is not surprising that so many HCW report burnout.^5^

The results of this study are consistent with prior studies demonstrating a high prevalence of insomnia in nursing and physicians caring for COVID-19 patients on the frontlines.^8^ Salari et al.^8^ report in a systematic review and meta-analysis on the increase of insomnia in frontline nurses near 35% and in frontline physicians near 42%, however they did not report on a sub-group analysis of insomnia in women. In the meta-analysis by Pappa et al.,^7^ insomnia prevalence in HCW during the COVID-19 pandemic was near 40%, and the authors did not report specifically the prevalence of insomnia in women frontline HCW. Our data is also in concert with a Trockel et al. ^5^ demonstrating sleep-related impairment as an occupational hazard associated with increased medical-related errors and burnout. In comparing our sample to the general U.S. physicians, we find higher rates of sleep disturbances and insomnia risk than the general population of practicing physicians.^22–24^ Our study findings of sleep disruption due to device usage are consistent with findings demonstrating increased nightly screen time in HCW and reports of “doomscrolling.” ^3,4^

There are many clinical implications of these findings. Frontline HCW involved in the direct care of COVID-19 patients report decreased sleep time, increased insomnia, nightmares, fears of safety, increased clinical workload, and concerns for their family, reportedly impacting their sleep. Healthcare workers are at high risk for the development of psychological distress and medical errors.^5,25^ The professional environment for healthcare workers has drastically changed with the pandemic and brought with it challenges related to increased workload, reduced protective equipment and resources, rapidly evolving protocols, relocation of intensive care settings, fear of viral transmission, and social isolation from supportive networks.^26,27^ Given this unfolding environment, findings of sleep disturbances of frontline HCW is not surprising.

The study findings are particularly concerning given the natural course of insomnia and potential for long term psychological impact, as seen in HCW during prior Severe Acute Respiratory Syndrome (SARS) outbreaks.^28–30^ In a recently reported longitudinal study on insomnia, nearly half (42%) of patients who had insomnia initially (as measured by the ISI) also had insomnia 5 years later, demonstrating a persistent course.^31^ Likewise, studies on the mental health of HCW active during the 2003 SARS epidemic suggest the psychological consequences may persist months to years after the epidemic ends.^30^ While noting long term impacts of mental health, it is also important to consider the implication of potential medical errors due to sleep loss by these HCW.^6^ Studies of sleep disorders in HCW during prior infectious outbreaks are in concert with our study findings, and they forecast the potential longitudinal psychological impact on the sleep health of our frontline healthcare colleagues.

Although social media platforms are widely utilized in 2020, a limitation of this study may be that recruitment through social media platforms may not be a representative sample of HCW in the U.S. Because the focus of our survey was on sleep, it is possible that HCW who were more likely to have a sleep disturbance completed the survey. Given the prevalence of sleep disorders in the general U.S. population of 8 to 19%,^32,33^ our sample does not appear to over-represent those with sleep disorders. There were not enough non-physician HCW in our sample to characterize the sleep quality for non-physician workers (e.g., physician’s assistant, nurse practitioner, nurse, pharmacist, allied health workers). We also asked subjects to complete the survey questions by reflecting on a time when they experienced the greatest clinical intensity and risk of COVID-19 transmission; this may have led to inconsistencies in our data. All data were self-reported, and we do not have objective measures of sleep duration and quality in this sample. We also do not have longitudinal data on our sample, limiting our ability to draw longitudinal conclusions. Furthermore, when utilizing social media data, it is difficult to capture the accurate outreach and link clicks; e.g., if an individual shares the link but not our original post, we cannot calculate the outreach and metrics. Therefore, the actual metrics may be higher than those we were capable of measuring.

## Conclusion

During the COVID-19 pandemic, 90% of HCW surveyed on social media reported poor sleep, with nearly half of respondents reporting moderate-severe insomnia and over half reporting burnout. Immediate interventions to improve HCW sleep and well-being, discourage nocturnal social media usage, and “doomscrolling” are needed to strengthen their ability to continuously meet daily demands of the COVID-19 pandemic. Considering that sleep is a modifiable factor of our physical and mental health, prioritizing it may help mitigate the health risks associated with the pandemic and potential longitudinal impacts on sleep and health.

## Data Availability

The data from this study is stored at The University of Chicago.

